# Predicting Individualized Outcomes for Deceased Kidney Donor Waitlisted Candidates and Recipients

**DOI:** 10.1101/2023.10.02.23296462

**Authors:** Igor Litvinovich, Yue-Harn Ng, Kelly Chong, Christos Argyropoulos, Yiliang Zhu

## Abstract

Kidney transplantation remains the optimal treatment for end-stage kidney disease (ESKD). However, the persistent disparity between the demand and supply of deceased donor (DD) kidneys underscores the need for better tools to assess transplant outcomes and donor kidney quality. The current Kidney Allocation System (KAS) relies on the Kidney Donor Risk Index (KDRI) to quantify DD kidney quality, yet it combines allograft failure and patient death into a single outcome, limiting its accuracy.

In this paper we present refined statistical models to predict post-transplantation risk, differentiating between allograft failure and patient death as competing risks. Using comprehensive data from the Organ Procurement and Transplantation Network/Scientific Registry of Transplant Recipient (OPTN/SRTR) for 2000-2017, our models incorporate biological and clinical factors instead of donor race, account for within-center clustering and between-center variation, and capture non-linear relationships between risk factors.

Our results reveal distinct risk factors for allograft failure and patient death. These models provide more personalized risk estimates tailored to donor kidney quality and recipient characteristics, aiding shared decision-making on kidney acceptance. Comparisons with the original KDRI demonstrate the superiority of our separate models, with improved predictability and reduced bias. Our approach eliminates the need to conflate allograft failure and patient death, leading to more accurate risk assessment and better-informed decisions regarding kidney offers.

In conclusion, our study underscores the importance of distinguishing between allograft failure and patient death in kidney transplant risk assessment. By offering more precise risk estimates, our models enhance the transparency and efficiency of kidney acceptance decisions, ultimately benefiting both transplant providers and candidates. We also provide a web-based tool to facilitate this process, promoting better outcomes in kidney transplantation.

**Key Points:**

1. Improved statistical models for kidney transplant risk assessment, separating the risks of allograft failure and patient death.
2. Models provide more personalized risk estimates, outperforming the existing Kidney Donor Risk Index (KDRI).
3. Models enhance transparency and accuracy in evaluating donor kidney quality, aiding both providers and candidates in decision-making.
4. Research improves the efficiency of kidney acceptance processes, leading to more successful transplants.

## Introduction

Kidney transplantation, even with donor kidneys of marginal quality, provides the best survival for patients with end-stage kidney disease (ESKD) compared to remaining on dialysis.^1–3^ The 2021 annual report of Organ Procurement and Transplantation Network/Scientific Registry of Transplant Recipient (OPTN/SRTR) showed the gap between the demand and supply of deceased donor (DD) kidneys remains wide with more candidates being placed on the wait list than candidates removed.^4^ Efforts to increase the number of kidney transplants include strategies to decrease kidney discards by utilizing older donor kidneys, donors with comorbidities including hypertension, diabetes as well as high-risk donors.^2, 5–10^ With the varied quality of deceased donor kidneys, there is an eminent need for tools that can help patients and providers more accurately estimate transplant outcomes along with donor kidney quality. Such data-driven tools would help transplant providers as well as transplant candidates to 1) best match a DD kidney with a candidate to achieve optimal transplant outcomes, and 2) aid in the decision of accepting a DD kidney.

The current kidney allocation system (KAS), implemented in December 2014, utilizes the kidney donor risk index (KDRI)^11^ to quantify the quality of DD kidneys based on probabilistic risk of kidney failure or patient death after transplant. A donor’s KDRI is converted into a percentile in reference to the KDRI distribution of all DD kidneys recovered from the prior year, and the percentile then becomes Kidney Donor Profile Index (KDPI). As a risk score; the higher the KDPI, the poorer the donor kidney quality. KDPI provides more granularity than the old Standard Criteria Donor/Expanded Criteria Donor classification. However, physicians and patients will still have to weigh the risk and benefit of each kidney offer and balance it with the risk of remaining on dialysis. More transparent and accurate evaluation of the multitudes of risks will likely aid in a shared decision making between the provider and the candidate on whether to accept a kidney offer.

Based on a comprehensive statistical model, the KDRI includes 10 donor factors identified through a statistical model of allograft failure or patient death whichever occurs first.^11^ It has an overall C statistic of 0.62, a measure of moderate level of model predictability, likely attributable to several limitations including combining two distinct outcomes, allograft failure (GF) and patient death, whichever occurs first, into a single one (Figure 1a). Patient death can be treated as a competing risk because a patient death not caused by kidney failure prevents the observation of kidney failure. Furthermore, the risk factors to and predictors of allograft failure and recipient death will likely differ both qualitatively and quantitatively. Thus, assuming that the two outcomes can be treated interchangeably will instill bias into the model predictions. Moreover, since the publication of the KDRI model in 2009, much has changed. Variables in the KDRI that were initially thought to be significant predictors of the outcomes have been called into question. A recent publication^12^ shows that donor race, a social construct, is at best a proxy of some associated biological and clinical factors. Upon fully incorporating these biological and clinical factors, the donor-race removed model predicts kidney transplant outcome equally well compared with the original KDRI model.^11^ The advent of the direct-acting antiviral (DAA) in the treatment of hepatitis C virus (HCV) has also changed the landscape of using HCV-positive kidney donors in kidney transplants.^13^ In view of these developments as well as the use of statistical techniques such as stratification by transplantation-year in Cox regression in the KDRI model,^11^ it is highly desirable to develop improved risk prediction models.

**Figure 1.**
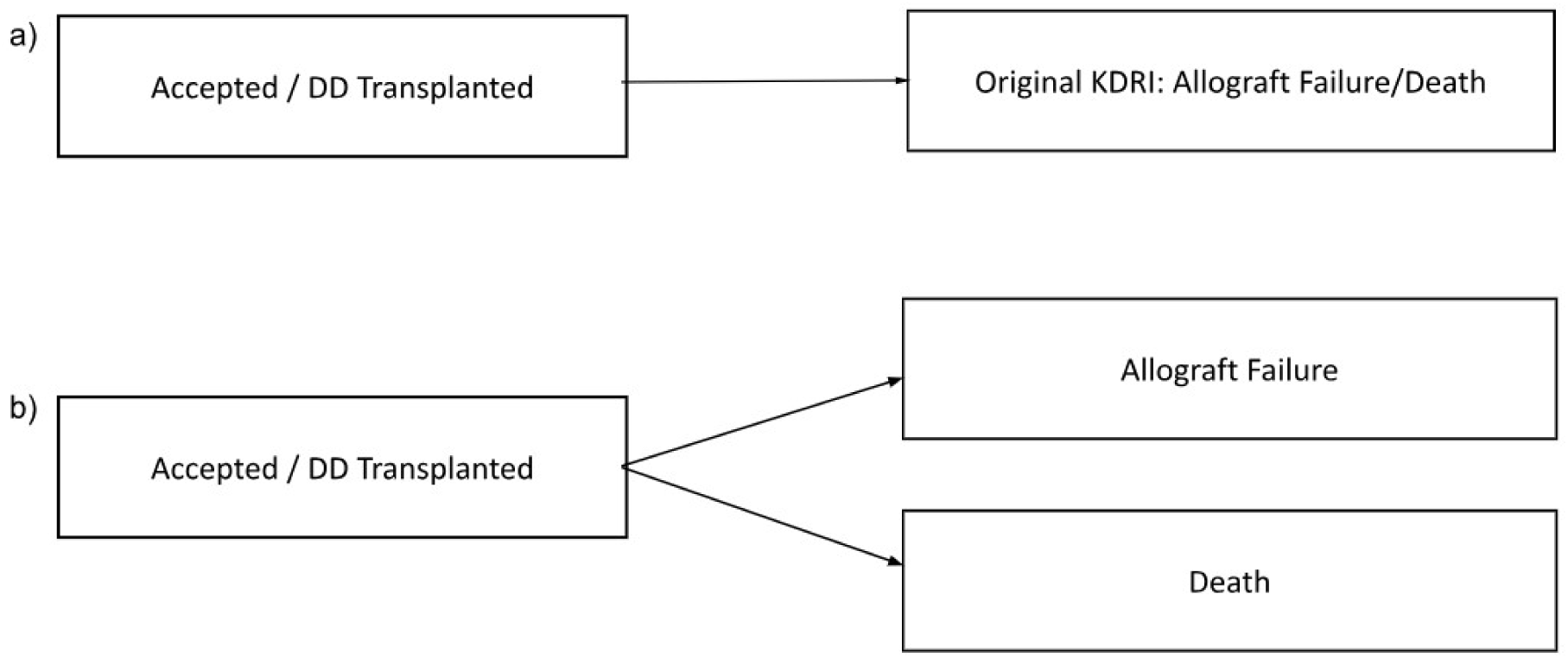
Scheme of modeling.

**Figure 2.**
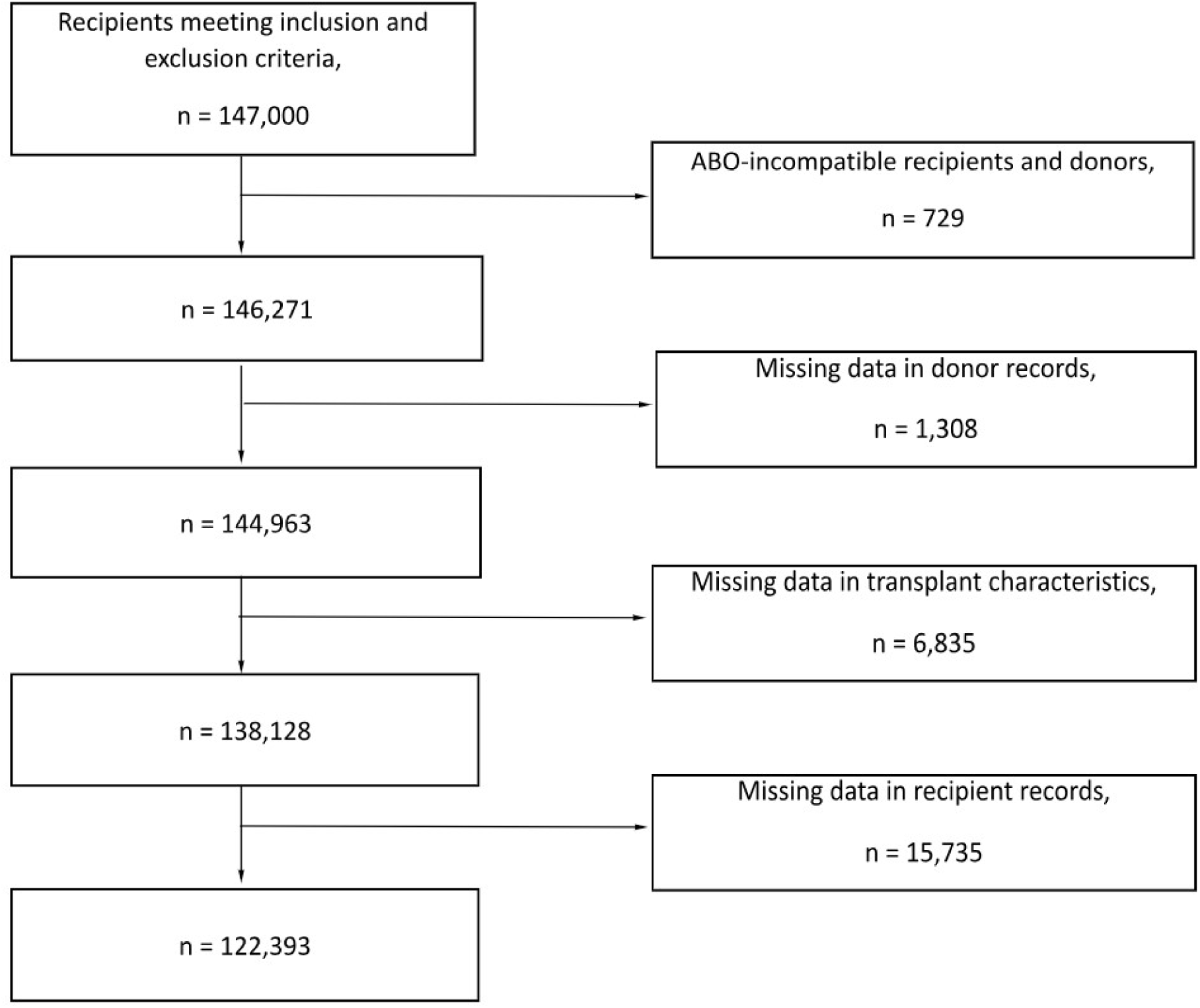
Flow chart of the sample selection process.

In this paper we present new models that predict the post-transplantation risk of allograft failure and the risk of patient death as a competing risk to allograft failure. Using 2000-2017 SRTR data, the models deploy refined options including removing donor race, better account for within-center clustering and between-center variation using both center characteristics and frailty, incorporating non-linear relationship between the risks and continuous risk factors such as donor weight and creatinine level. Our objective is to provide more personalized risk estimates of the distinct risks of allograft failure or patient death specific to donor kidney quality and recipient characteristics. Such personalized risk estimates will better inform a shared-decision making on accepting a DD kidney.

## Materials and Methods

We obtained data from SRTR of all transplanted kidneys in the US between January 1, 2000 and December 31, 2017. The SRTR data includes characteristics of donors and wait-listed candidates, as well as transplantation factors for transplant kidneys. For this analysis, we included 1) adult transplant candidates of 18 years or older; 2) kidney transplantation only with no prior solid organ transplants; 3) deceased donor kidney transplant. We excluded living donor transplants, multi-organ transplant recipients, and recipients with any prior solid organ transplants. The final dataset included a total of 147,000 adult recipients of first-time kidney-only transplants with deceased donors.

We developed a Cox survival model for post-transplant allograft failure while treating patient death as a competing risk outcome with a separate survival model. If allograft failure was observed prior to patient death, allograft failure is the primary outcome; if patient death was observed prior to a recorded allograft failure, death is treated as a competing risk that prevents the observation of allograft failure. Instead of using donor race as a proxy, we used a number of biological and clinical factors to replace donor race in the model, as supported by the latest evidence.^12^ We explored and included a large number of donor and recipient characters as well as transplantation factors as potential predictors. Our selection of predictors was in part informed by the original KDRI model^11^ and determined by both clinical relevance and statistical model selection criteria. In the original KDRI model,^11^ continuous donor factors were made piece-wise linear over selected intervals when a non-linear relationship with the hazard was supported by data. We adopted penalized spline functions to identify a non-linear relationship and used the intervals suggested by the penalized spline functions to construct a piecewise linear function to approximate the non-linear relationship. We used transplantation year as strata to account for year-to-year variation in the hazard functions for allograft failure and patient death when the assumption of proportionality of hazards by year of transplantation may be violated. We also used frailty (random effects) to account for within-center clustering and between-center variation in transplant outcomes. These options are in contrast with the use of transplantation center, recipient age, and recipient status of diabetes as stratification factors in Rao et al.^11^ We also used false discovery rate (FDR) adjustment for p-values for individual predictors in the model to control the familial type I error to be at the nominal level 0.01.

For variable selection and assessing model fitting we used statistical criteria such as the Akaike information criterion (AIC) and statistical significance. For overall model predictability we used the concordance index (C-statistic) and the area under the curve (AUC) of the Receiver Operating Characteristic (ROC) curve. A ROC curve in the present case of the survival model shows the relationship between true positive prediction (sensitivity) and false negative (one minus specificity) using hazard-based predictors at each point in time with an observed event, hence is time-dependent. The AUC measures the overall model performance when using all possible hazard thresholds for prediction at a given time point. We estimated AUCs using R package survAUC.^14–15^ To implement the time-dependent ROC, we first randomly selected 40,000 records as the training sample to fit the respective models for allograft failure and patient death, and then used the fitted models to predict outcomes for the remaining (validation) sample to report AUC at each month post-transplantation over the 18-year study period. We replicated this process 10 times and calculated the average AUC of the replications. We used R version 4.2.1 for all analyses.

This study was approved by the Institutional Review Board at the University of New Mexico.

## Results

Out of the 147,000 adult first-time kidney-only transplant recipients, we excluded 729 patients who were ABO-incompatible. Additionally, we excluded 23,878 unique transplantation records, of which 1,308 recipients with missing donor characteristics, 6,835 recipients with missing transplant characteristics and 15,735 recipients with missing patient characteristics, which were required in our models. A comparison of the patients and donors with missing data is reported in Supplement Table 1. The final dataset included 122,393 unique transplants, of which 18,897 (15.4%) had allograft failure and 20,802 (17.0%) patients died without a diagnosis of allograft failure in the follow-up period up to December 31, 2017. We identified allograft failures and the failure date based on the record of allograft failure diagnosis, returning to dialysis, or re-transplant, whichever occurred first.

In Table 1, we summarize donor and recipient characters as well as transplantation factors. Black donors were more likely to be younger, have cerebrovascular accident (CVA) death, diabetes, hypertension, and higher levels of serum creatinine, and less likely to have circulatory death (DCD); and these differences were statistically significant. Further, Black donors were more likely to be used for en bloc or double transplantation than non-Black donors (4.3% vs. 2.8%), and have slightly longer average cold ischemic time (18.3 ± 9.4 vs. 17.8± 9.0). We note that a Black donor was more likely to be transplanted to a Black recipient than a White recipient (50.4% vs 42.7%), and a non-Black donor was more likely to be transplanted to a White recipient than a Black recipient (60.7% vs 30.5%). A Black donor was more likely to be transplanted in a patient on dialysis for a longer period (wait-listed longer). Finally, a Black donor was more likely to be transplanted into a recipient with a higher calculated Panel Reactive Antibodies (cPRA) or with cardiovascular disease.

**Table 1.**
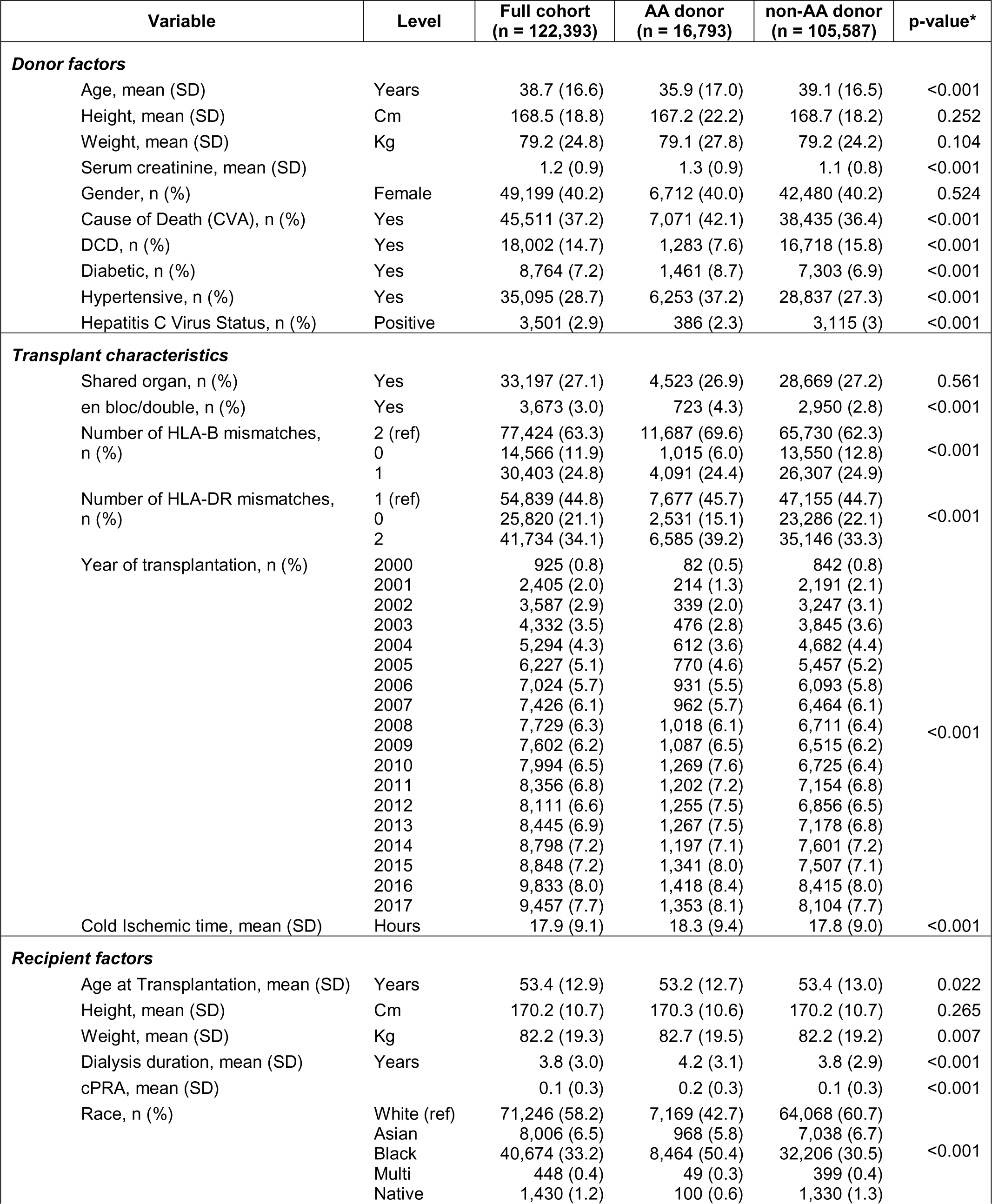
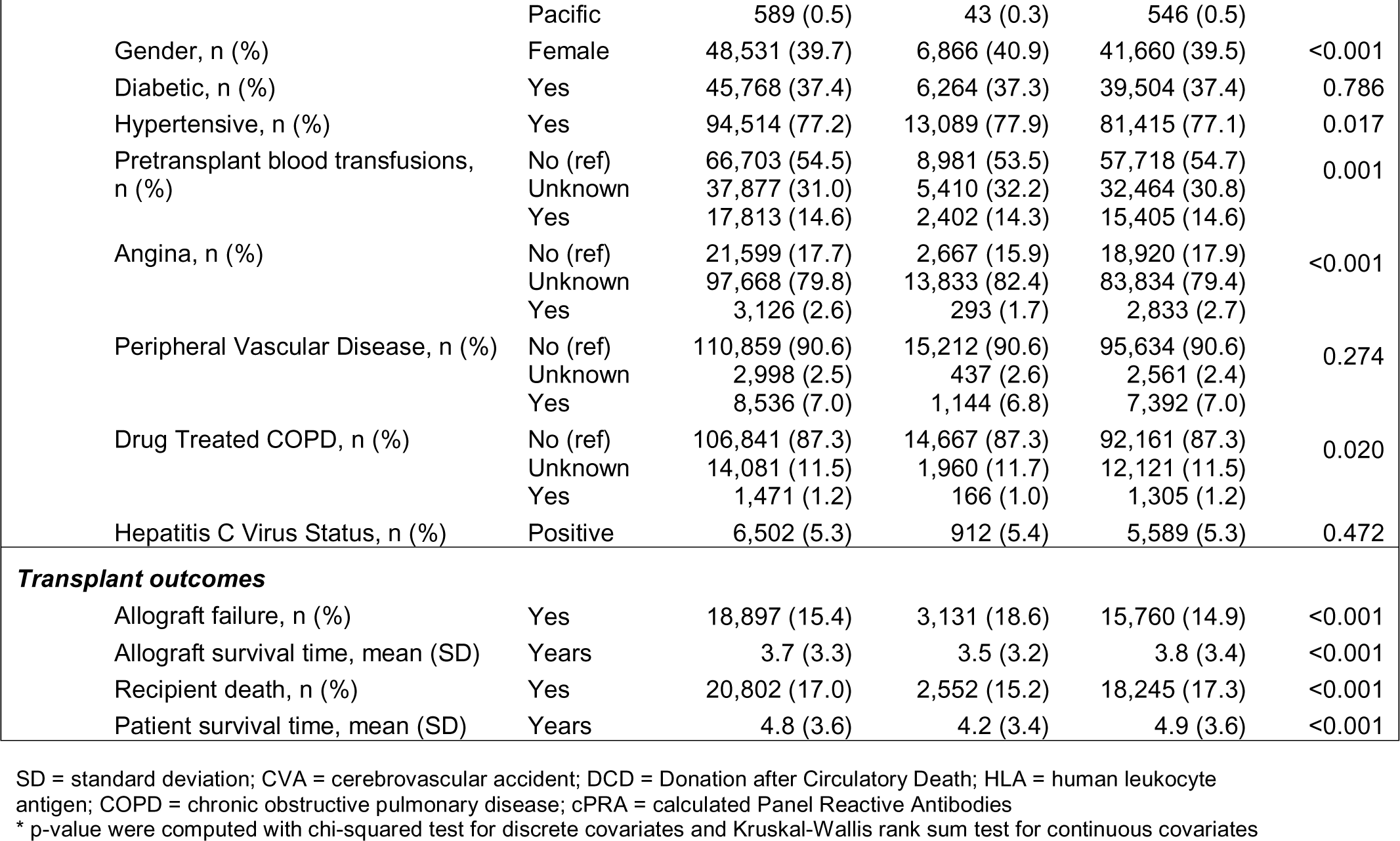
Data Summary Table.

Table 2 summarizes the Cox survival models for allograft failure and patient death, respectively, for first-time solid organ recipients of deceased donor kidneys. In the risk model for death, only death without prior allograft failures was included as outcome. Separating patient deaths as a competing risk outcome to allograft failure led to a modified KDRI with two components, KDRI_a for allograft failure and KDRI_d for patient death on the basis of the respective Cox regression model.

**Table 2.**
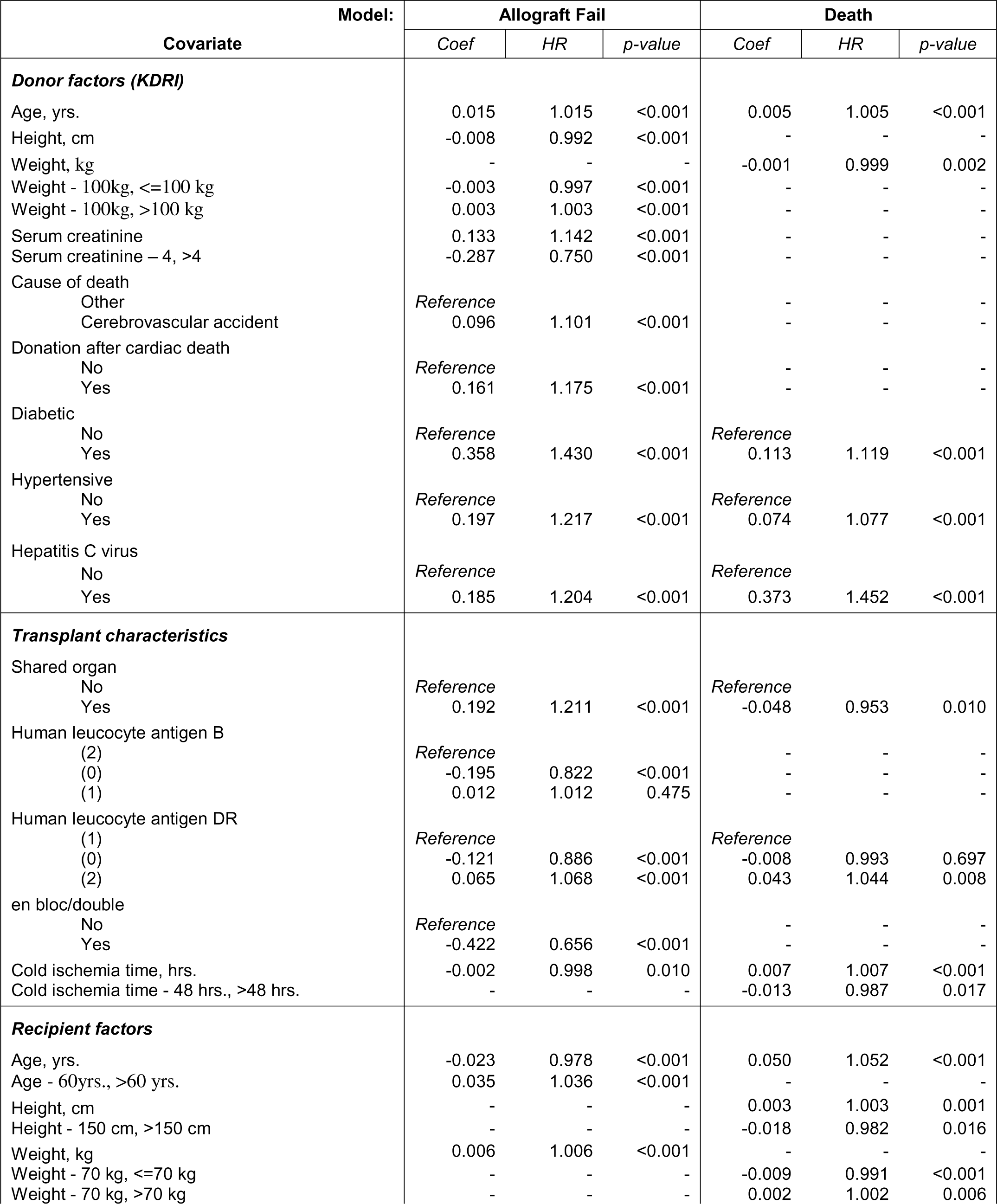
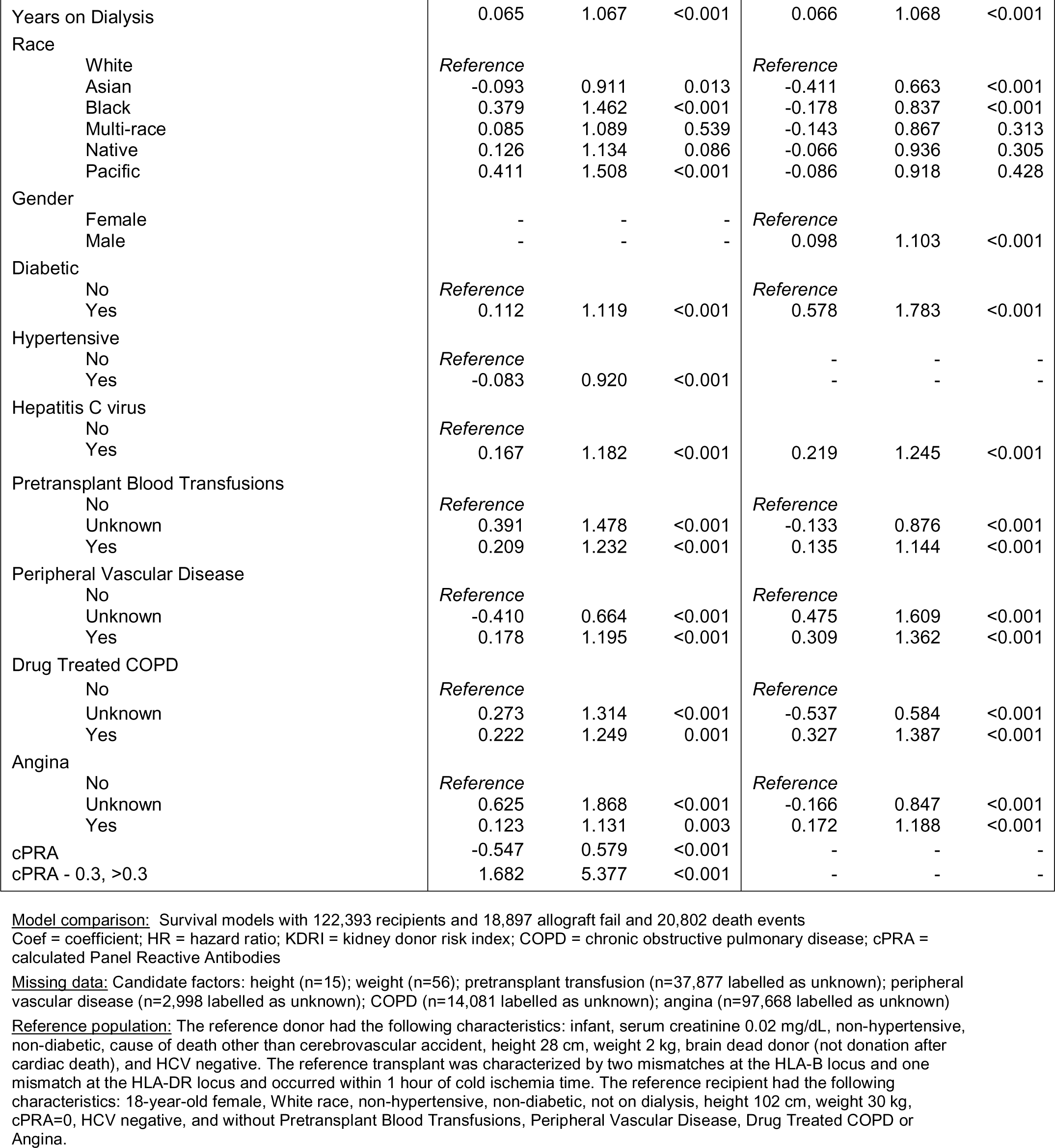
Survival models for allograft failure or patient deaths.

| Covariate | Model: | Allograft Fail |  |  | Death |  |  |
| --- | --- | --- | --- | --- | --- | --- | --- |
|  |  | Coef | HR | p-value | Coef | HR | p-value |
| **Donor factors (KDRI)** |  |  |  |  |  |  |  |
| Age, yrs. |  | 0.015 | 1.015 | <0.001 | 0.005 | 1.005 | <0.001 |
| Height, cm |  | -0.008 | 0.992 | <0.001 | - | - | - |
| Weight, kg |  | - | - | - | -0.001 | 0.999 | 0.002 |
| Weight - 100kg, <=100 kg |  | -0.003 | 0.997 | <0.001 | - | - | - |
| Weight - 100kg, >100 kg |  | 0.003 | 1.003 | <0.001 | - | - | - |
| Serum creatinine |  | 0.133 | 1.142 | <0.001 | - | - | - |
| Serum creatinine – 4, >4 |  | -0.287 | 0.750 | <0.001 | - | - | - |
| Cause of death |  |  |  |  |  |  |  |
| Other |  | Reference |  |  | - | - | - |
| Cerebrovascular accident |  | 0.096 | 1.101 | <0.001 | - | - | - |
| Donation after cardiac death |  |  |  |  |  |  |  |
| No |  | Reference |  |  | - | - | - |
| Yes |  | 0.161 | 1.175 | <0.001 | - | - | - |
| Diabetic |  |  |  |  |  |  |  |
| No |  | Reference |  |  | Reference |  |  |
| Yes |  | 0.358 | 1.430 | <0.001 | 0.113 | 1.119 | <0.001 |
| Hypertensive |  |  |  |  |  |  |  |
| No |  | Reference |  |  | Reference |  |  |
| Yes |  | 0.197 | 1.217 | <0.001 | 0.074 | 1.077 | <0.001 |
| Hepatitis C virus |  |  |  |  |  |  |  |
| No |  | Reference |  |  | Reference |  |  |
| Yes |  | 0.185 | 1.204 | <0.001 | 0.373 | 1.452 | <0.001 |
| **Transplant characteristics** |  |  |  |  |  |  |  |
| Shared organ |  |  |  |  |  |  |  |
| No |  | Reference |  |  | Reference |  |  |
| Yes |  | 0.192 | 1.211 | <0.001 | -0.048 | 0.953 | 0.010 |
| Human leucocyte antigen B |  |  |  |  |  |  |  |
| (2) |  | Reference |  |  | - | - | - |
| (0) |  | -0.195 | 0.822 | <0.001 | - | - | - |
| (1) |  | 0.012 | 1.012 | 0.475 | - | - | - |
| Human leucocyte antigen DR |  |  |  |  |  |  |  |
| (1) |  | Reference |  |  | Reference |  |  |
| (0) |  | -0.121 | 0.886 | <0.001 | -0.008 | 0.993 | 0.697 |
| (2) |  | 0.065 | 1.068 | <0.001 | 0.043 | 1.044 | 0.008 |
| en bloc/double |  |  |  |  |  |  |  |
| No |  | Reference |  |  | - | - | - |
| Yes |  | -0.422 | 0.656 | <0.001 | - | - | - |
| Cold ischemia time, hrs. |  | -0.002 | 0.998 | 0.010 | 0.007 | 1.007 | <0.001 |
| Cold ischemia time - 48 hrs., >48 hrs. |  | - | - | - | -0.013 | 0.987 | 0.017 |
| **Recipient factors** |  |  |  |  |  |  |  |
| Age, yrs. |  | -0.023 | 0.978 | <0.001 | 0.050 | 1.052 | <0.001 |
| Age - 60yrs., >60 yrs. |  | 0.035 | 1.036 | <0.001 | - | - | - |
| Height, cm |  | - | - | - | 0.003 | 1.003 | 0.001 |
| Height - 150 cm, >150 cm |  | - | - | - | -0.018 | 0.982 | 0.016 |
| Weight, kg |  | 0.006 | 1.006 | <0.001 | - | - | - |
| Weight - 70 kg, <=70 kg |  | - | - | - | -0.009 | 0.991 | <0.001 |
| Weight - 70 kg, >70 kg |  | - | - | - | 0.002 | 1.002 | 0.006 |

|  |  |  |  |  |  |  |
| --- | --- | --- | --- | --- | --- | --- |
| Years on Dialysis | 0.065 | 1.067 | <0.001 | 0.066 | 1.068 | <0.001 |
| Race |  |  |  |  |  |  |
| White | *Reference* |  |  | *Reference* |  |  |
| Asian | -0.093 | 0.911 | 0.013 | -0.411 | 0.663 | <0.001 |
| Black | 0.379 | 1.462 | <0.001 | -0.178 | 0.837 | <0.001 |
| Multi-race | 0.085 | 1.089 | 0.539 | -0.143 | 0.867 | 0.313 |
| Native | 0.126 | 1.134 | 0.086 | -0.066 | 0.936 | 0.305 |
| Pacific | 0.411 | 1.508 | <0.001 | -0.086 | 0.918 | 0.428 |
| Gender |  |  |  |  |  |  |
| Female | - | - | - | *Reference* |  |  |
| Male | - | - | - | 0.098 | 1.103 | <0.001 |
| Diabetic |  |  |  |  |  |  |
| No | *Reference* |  |  | *Reference* |  |  |
| Yes | 0.112 | 1.119 | <0.001 | 0.578 | 1.783 | <0.001 |
| Hypertensive |  |  |  |  |  |  |
| No | *Reference* |  |  | - | - | - |
| Yes | -0.083 | 0.920 | <0.001 | - | - | - |
| Hepatitis C virus |  |  |  |  |  |  |
| No | *Reference* |  |  |  |  |  |
| Yes | 0.167 | 1.182 | <0.001 | 0.219 | 1.245 | <0.001 |
| Pretransplant Blood Transfusions |  |  |  |  |  |  |
| No | *Reference* |  |  | *Reference* |  |  |
| Unknown | 0.391 | 1.478 | <0.001 | -0.133 | 0.876 | <0.001 |
| Yes | 0.209 | 1.232 | <0.001 | 0.135 | 1.144 | <0.001 |
| Peripheral Vascular Disease |  |  |  |  |  |  |
| No | *Reference* |  |  | *Reference* |  |  |
| Unknown | -0.410 | 0.664 | <0.001 | 0.475 | 1.609 | <0.001 |
| Yes | 0.178 | 1.195 | <0.001 | 0.309 | 1.362 | <0.001 |
| Drug Treated COPD |  |  |  |  |  |  |
| No | *Reference* |  |  | *Reference* |  |  |
| Unknown | 0.273 | 1.314 | <0.001 | -0.537 | 0.584 | <0.001 |
| Yes | 0.222 | 1.249 | 0.001 | 0.327 | 1.387 | <0.001 |
| Angina |  |  |  |  |  |  |
| No | *Reference* |  |  | *Reference* |  |  |
| Unknown | 0.625 | 1.868 | <0.001 | -0.166 | 0.847 | <0.001 |
| Yes | 0.123 | 1.131 | 0.003 | 0.172 | 1.188 | <0.001 |
| cPRA | -0.547 | 0.579 | <0.001 | - | - | - |
| cPRA - 0.3, >0.3 | 1.682 | 5.377 | <0.001 | - | - | - |

Model comparison: Survival models with 122,393 recipients and 18,897 allograft fail and 20,802 death events
Coef = coefficient; HR = hazard ratio; KDRI = kidney donor risk index; COPD = chronic obstructive pulmonary disease; cPRA = calculated Panel Reactive Antibodies
Missing data: Candidate factors: height (n=15); weight (n=56); pretransplant transfusion (n=37,877 labelled as unknown); peripheral vascular disease (n=2,998 labelled as unknown); COPD (n=14,081 labelled as unknown); angina (n=97,668 labelled as unknown)
Reference population: The reference donor had the following characteristics: infant, serum creatinine 0.02 mg/dL, non-hypertensive, non-diabetic, cause of death other than cerebrovascular accident, height 28 cm, weight 2 kg, brain dead donor (not donation after cardiac death), and HCV negative. The reference transplant was characterized by two mismatches at the HLA-B locus and one mismatch at the HLA-DR locus and occurred within 1 hour of cold ischemia time. The reference recipient had the following characteristics: 18-year-old female, White race, non-hypertensive, non-diabetic, not on dialysis, height 102 cm, weight 30 kg, cPRA=0, HCV negative, and without Pretransplant Blood Transfusions, Peripheral Vascular Disease, Drug Treated COPD or Angina.

The allograft failure model included most donor factors that are components of the original KDRI model,^11^ resulting in KDRI_a as follows:

KDRI_a = 0.015*Age - 0.008*Height - 0.003*(Weight - 100)*I(Weight <= 100) + 0.003*(Weight - 100)*I(Weight > 100) + 0.133*Creatinine - 0.287*(Creatinine - 4)*I(Creatinine > 4) + 0.096*CVA + 0.161*DCD + 0.358*Diabetes + 0.197*Hypertension + 0.185*I(Donor HCV positive).

In the equation above, I(.) is an indicator function which is equal to 1 when the condition(s) inside the parentheses holds true, and 0 otherwise. CVA, DCD, diabetes, hypertension, and HCV are dichotomized indicators of the presence of the corresponding condition.

In the final patient death model (Table 2) only donor factors that were significant predictors of death risk were age, height, gender, diabetes, hypertension, and HCV status as significant risk factors. As a result, KDRI_d is given by the follow: KDRI_d = 0.005*Age - 0.001*Weight + 0.113*Diabetes + 0.074*Hypertension + 0.373*I(Donor HCV positive).

For comparison, we plotted the original KDRI against KDRI_a and KDRI_d for Black donors and non-Black donors separately (Figure 3). Note that the original KDRI, the new KDRI_a and KDRI_d are all a function of hazard ratio relative to a hypothetical reference donor population. For KDRI, the reference population please see the footnotes of Table 2. First, Figure 3 shows that the distribution is different for KDRI_a and KDRI_d. KDRI_a has a much wider range than KDRI_d besides different reference populations. By lump allograft failure and patient death into a composite outcome, the differences in relative risks for allograft failure and patient death are masked. Secondly, the distributions of KDRI_a and KDRI_d are comparable in both shape and scale between Black and non-Black donors, suggesting a limited role the Black donor race may play in the KDRI profile.

**Figure 3.**
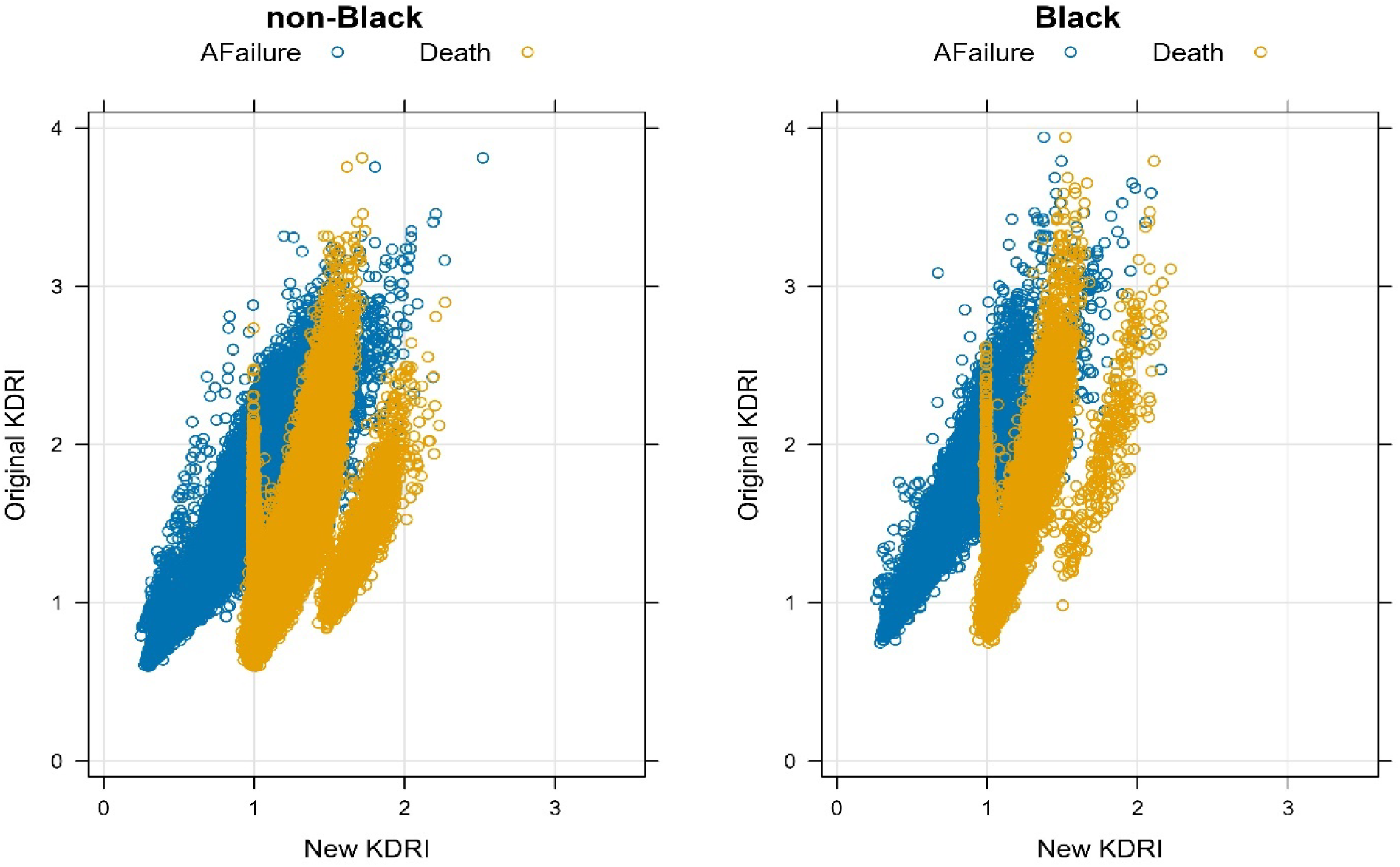
KDRI comparison.

**Figure 4.**
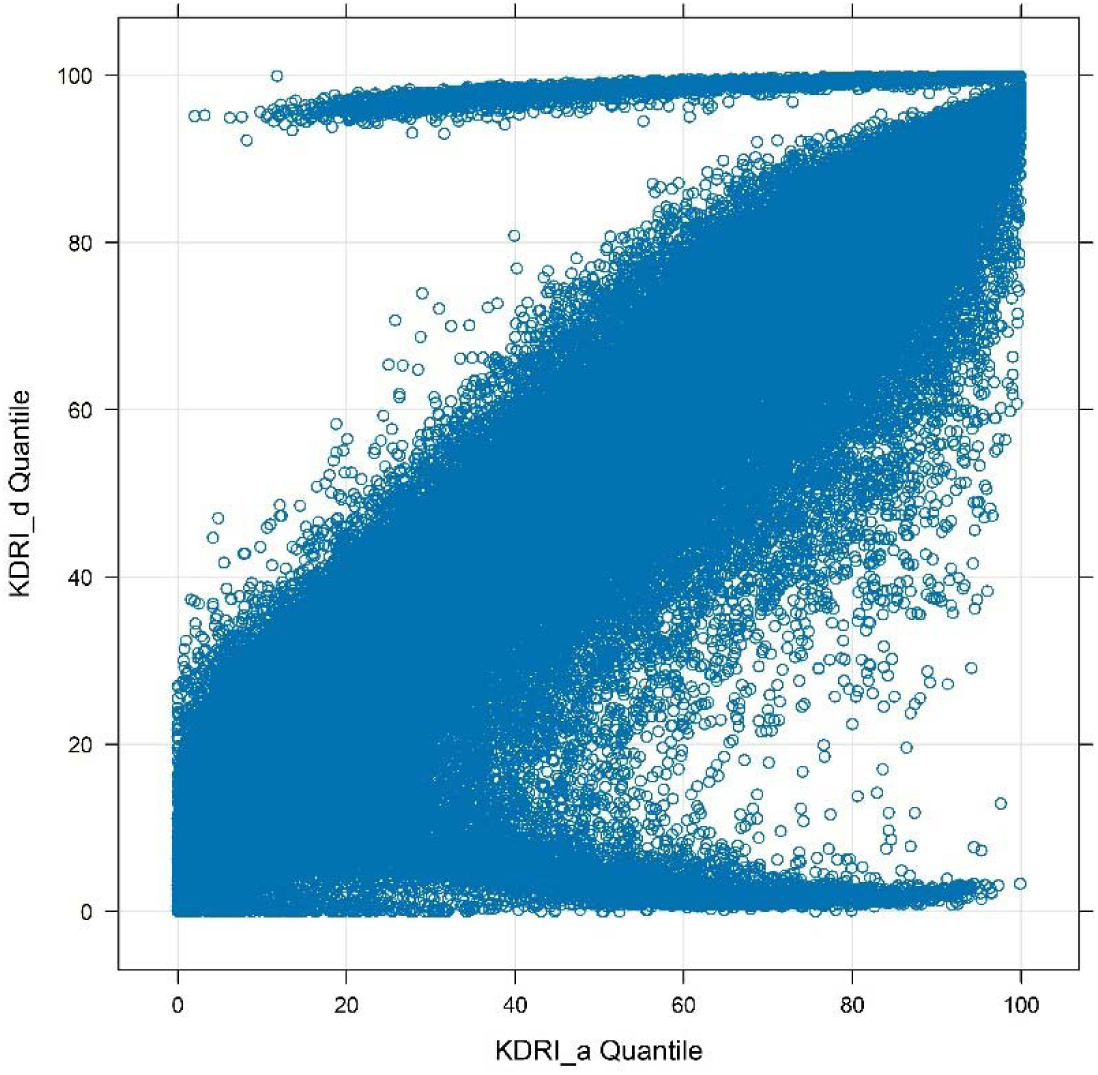
KDRI_a and KDRI_b comparison.

The differences in KDRI_a and KDRI_d are clearly seen in the donor factors that contribute to the models (Table 2). Whereas donor weight, serum creatinine level, cerebrovascular death, and donation after cardiac death are significant contributing factors to KDRI_a, they are not to KDRI_d. Moreover, the impacts of diabetes and hypertension of a donor on increased risk of allograft failure were three times as greater as those on the increased risk of patient death. The risks of allograft failure and patient death also differ in transplant factors. More specifically, the models and resultant KDRI_a and KDRI_d suggest that en bloc/double kidneys transplants were associated with reduced risks of allograft failure, and no effect on patient death.

Finally, risks of allograft failure and patient death also differ in recipient characteristics. Increasing recipient age was associated with decreased risk of allograft failure but increased risk of patient death. It is noteworthy that Black recipients had (41%) higher risk (hazard ratio) of allograft failure but (15%) lower risk of patient death than White recipients on average. The risk of death was 77% greater among recipients with diabetes than recipients without diabetes, and 5 times of the increased risk of allograft failure associated with DM. We also note that a unit increase in cPRA was associated with 1.6 times increase in hazard of allograft failure, but was not associated with the risk of patient death.

Figure 5 displays the Kaplan-Meier survival curves based on the Cox models fitted to allograft failure, patient death, or the combined outcomes (the original model^11^), respectively. While the data-based patient death depicts a median survival of 13.1 years and median survival of allograft 16.6 years, the combined outcome model^11^ depicts a median survival of only 9.2 years, without differentiating the risks of allograft failure and patient death. It is interesting to note that while the allograft survival probability decreases approximately linearly, the risk of patient death accelerates with survival time.

**Figure 5.**
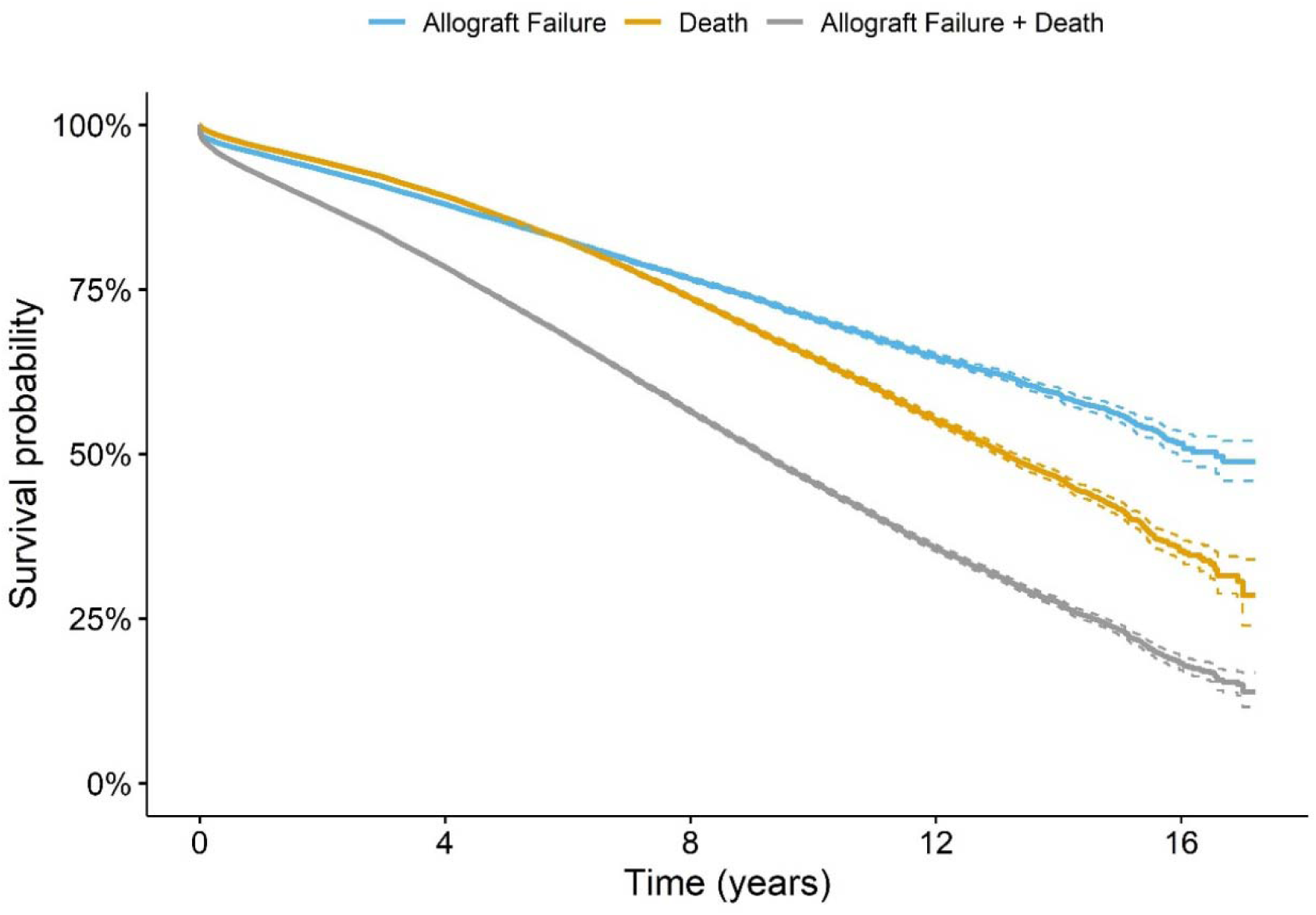
Kaplan-Meier estimates.

The time-dependent AUCs (Figure 6) show clearly that the model with the combined outcome of allograft failure and patient death performed poorly up to 10 years after transplantation in comparison with the separate models for allograft failure and patient death. In general, the predictive power of these models improved over time, especially for patient death. The AUC for allograft failure stayed above 0.67, and only improved over time slightly, indicating the complexity of the associated risk. The AUC of the combined outcome was lower than both of allograft failure and patient death up to year 11 post-transplant, it steadily increased afterwards as it was increasingly dominated perhaps by patient deaths. Overall the models for allograft failure and patient death have an average concordance index greater than that of the combined outcome model as proposed by Rao et al (C = 0.68, C = 0.72 vs. C = 0.62, respectively). Additionally, the computed false discovery rate (FDR) was very small for the GF model and was close to zero for the Death model (p-value threshold was 0.01). Our separate risk models demonstrate marked improvements in predicting transplantation risks, compared with models based on the combined outcome of allograft failure and patient death (Rao et al^11^ and Chong et al^12^). Notably, our models also removed Black donor race as a predictor, adjusted for non-linear relationship between the outcomes and continuous predictors using spline functions, and accounted for between-center variation using frailty.

**Figure 6.**
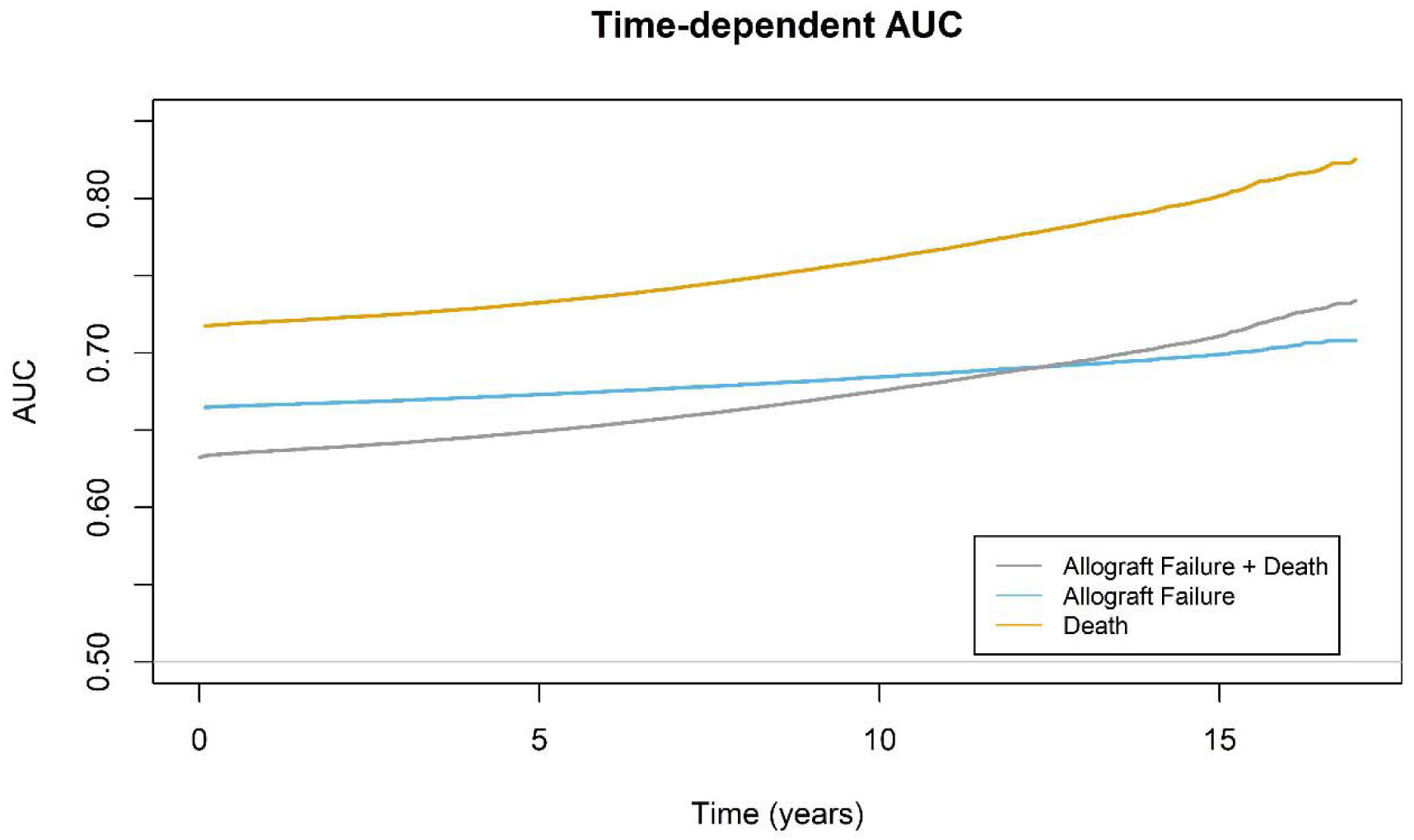
Time-dependent AUC.

## Discussion

We have demonstrated that the risk of allograft failure and the risk of patient death in the absence of allograft failure are distinct with respect to donor and recipient characteristics as well as transplant factors such as cold ischemic time and donor-recipient sex pairing. These distinct features offer strong clinical evidence supporting separate consideration of the two risks. As a result, we developed two Cox survival models to predict post-transplant allograft failure and patient death, and based on this developed one kidney donor risk sub-index for each.

The donor risk index for allograft failure retains most donor risk factors that are in the original KDRI, but only a subset pertains to the donor risk index for patient death. Notably, both sub-indices excluded most donor factors and transplant characteristics from the original model^11^ have an effect on allograft survival, but only some of them have an effect on patient survival.

We used model-based spline functions to address non-linear relationships between predictors and hazard. We found that the donor’s weight has a different effect on allograft survival at 100 kg cut-off point; the recipient’s weight has a different effect on patient survival at 70 kg cut-off point.

Using statistical methods and criteria in predictor selection and model building, we created models for allograft failure and patient death that outperformed the original model.^11^ Modified kidney donor risk indices were suggested to assess specific risks for allograft failure and patient death.

In conclusion, we have shown that the donor’s effect on post-transplant allograft and patient survival should be considered separately, and we created a web-based tool^16^ to facilitate the shared patient-provider decision to accept a kidney offer. This tool improves the transparency and efficiency of the kidney acceptance process based on more precise prediction of risks calculated specific to the donor-recipient pair instead of the population average.

## Supporting information

Table S1. Comparison of donor and recipient characteristics with and without missing data

## Data Availability

Comprehensive data from the Organ Procurement and Transplantation Network/Scientific Registry of Transplant Recipient for 2000-2017 were used.

## Author’s Contribution

Research idea and study design: YZ, CA, IL and YN; statistical analysis: IL and YZ; data analysis/interpretation/drafting and revision of the manuscript: KC, IL, YZ, CA and YN. Each author contributed important intellectual content during manuscript drafting or revision and agrees to be personally accountable for the individual’s own contributions and to ensure that questions pertaining to the accuracy or integrity of any portion of the work, even one in which the author was not directly involved, are appropriately investigated and resolved, including with documentation in the literature if appropriate.

## References

1. Wolfe R, Ashby V, Milford E, Ojo A, Ettenger R, Agodoa L et al. Comparison of Mortality in All Patients on Dialysis, Patient on Dialysis Awaiting Transplantation, and Recipients of a First Cadaveric Transplant. New England Journal of Medicine 1999; 341: 1725–1730.

2. Ojo AO, Hanson JA, Meier-Kriesche H-U, Okechukwu CN, Wolfe RA, Leichtman AB, et al. Survival in Recipients of Marginal Cadaveric Donor Kidneys Compared with Other Recipients and Wait-Listed Transplant Candidates. Journal of the American Society of Nephrology 2001; 12(3): 589–597.

3. Kaballo MA, Canney M, O’Kelly P, Williams Y, O’Seaghdha CM, Conlon PJ. A comparative analysis of survival of patients on dialysis and after kidney transplantation. Clinical Kidney Journal 2018; 11(3): 389–393.

4. Optn. A Guide to Calculating and Interpreting the Kidney Donor Profile Index (KDPI). 2018; Available at: https://optn.transplant.hrsa.gov/data/allocation-calculators/kdpi-calculator/.

5. Merion RM, Ashby VB, Wolfe RA, et al. Deceased-donor characteristics and the survival benefit of kidney transplantation. JAMA 2005; 294(21): 2726–2733.

6. Metzger RA, Delmonico FL, Feng S, Port FK, Wynn JJ, Merion RM. Expanded criteria donors for kidney transplantation. American Journal of Transplantation 2003; 3(1): 114–125.

7. Maggiore Umberto U. The marginal kidney donor. Current Opinion in Organ Transplantation; 19(4): 372–380.

8. Pérez-Sáez MJ, Montero N, Redondo-Pachón D, Crespo M, Pascual J. Strategies for an Expanded Use of Kidneys from Elderly Donors. Transplantation 2017; 101(4): 727–745.

9. Port FK, Bragg-Gresham JL, Metzger RA, Dykstra DM, Gillespie BW, Young EW et al. Donor characteristics associated with reduced graft survival: an approach to expanding the pool of kidney donors1. Transplantation 2002; 74(9): 1281–1286.

10. Sharma N, Mahajan A, Qazi YA. Marginal kidney transplantation: the road less traveled. Current Opinion in Organ Transplantation 2019; 24(1): 92–96.

11. Rao PS, Schaubel DE, Guidinger MK, Andreoni KA, Wolfe RA, Merion RM et al. A comprehensive risk quantification score for deceased donor kidneys: the kidney donor risk index. Transplantation 2009; 88(2): 231–236.

12. Chong K, Litvinovich I, Chen SS, Zhu Y, Argyropoulos C, Ng Y-H. Reconsidering Donor Race in Predicting Allograft and Patient Survival Among Kidney Transplant Recipients. Kidney360. 2021;2(11): 1831–1835.

13. Kling CE, Perkins JD, Sibulesky L. Three-year follow-up of aviremic hepatitis C– positive kidneys. American Journal of Transplantation 2019; 19(11): 3212–3213.

14. Chambless LE, Diao G: Estimation of time-dependent area under the ROC curve for long-term risk prediction. Stat Med 25: 3474–3486, 2006. Available at: 10.1002/sim.2299.

15. Potapov S, Adler W, Schmid M: Package ‘survAUC’, 2015. Available at: https://cran.r-project.org/web/packages/survAUC/survAUC.pdf. Accessed September 28, 2021.

16. Kidney Risk Calculator. Available at https://kcalculator.shinyapps.io/kidneycalc/.

