## Supplementary material for "Predicting Individualized Outcomes for Deceased Kidney Donor Waitlisted Candidates and Recipients": Table S1. Comparison of donor and recipient characteristics with and without missing data

| **Variable** | **Level** | **Full cohort***  **(n = 146,271)** | **Missing data**  **(n = 23,878)** | **non-Missing data**  **(n = 122,393)** | **p-value**** |
| --- | --- | --- | --- | --- | --- |
| ***Donor factors*** |  |  |  |  |  |
| Age, mean (SD) | Years | 38.6 (16.7) | 38.2 (17.0) | 38.7 (16.6) | <0.001 |
| Height, mean (SD) | Cm | 168.5 (18.7) | 168.6 (18.1) | 168.5 (18.8) | 0.404 |
| not reported, n (%) |  | 5 (0.0) | 5 (0.0) | 0 (0.0) |  |
| Weight, mean (SD) | Kg | 78.7 (24.6) | 76.2 (23.4) | 79.2 (24.8) | <0.001 |
| Serum creatinine, mean (SD) |  | 1.2 (1.0) | 1.1 (1.0) | 1.2 (1.0) | 0.002 |
| not reported, n (%) |  | 88 (0.1) | 88 (0.4) | 0 (0) |  |
| Gender, n (%) | Female | 58,804 (40.2) | 9,605 (40.2) | 49,199 (40.2) | 0.942 |
| Cause of Death (CVA), n (%) | Yes | 56,348 (38.5) | 10,837 (45.4) | 45,511 (37.2) | <0.001 |
| not reported, n (%) |  | 3 (0.0) | 3 (0.0) | 0 (0.0) |  |
| DCD, n (%) | Yes | 19,438 (13.3) | 1,436 (6.0) | 18,002 (14.7) | <0.001 |
| Diabetic, n (%) | Yes | 9,949 (6.8) | 1,185 (5.0) | 8,764 (7.2) | <0.001 |
| not reported, n (%) |  | 699 (0.5) | 699 (2.9) | 0 (0.0) |  |
| Hypertensive, n (%) | Yes | 40,717 (27.8) | 5,622 (23.5) | 35,095 (28.7) | <0.001 |
| not reported, n (%) |  | 923 (0.6) | 923 (3.9) | 0 (0.0) |  |
| Hepatitis C Virus Status, n (%) | Positive | 3,990 (2.7) | 489 (2.1) | 3,501 (2.9) | <0.001 |
| ***Transplant characteristics*** |  |  |  |  |  |
| Shared organ, n (%) | Yes | 39,493 (27.0) | 6,296 (26.4) | 33,197 (27.1) | 0.017 |
| en bloc/double, n (%) | Yes | 4,442 (3.0) | 769 (3.2) | 3,673 (3.0) | 0.074 |
| Number of HLA-B mismatches,  n (%) | 2 (ref) | 90,529 (61.9) | 13,105 (54.9) | 77,424 (63.3) | <0.001 |
|  | 0 | 18,185 (12.4) | 3,619 (15.2) | 14,566 (11.9) |  |
|  | 1 | 37,546 (25.7) | 7,143 (29.9) | 30,403 (24.8) |  |
| not reported, n (%) |  | 11 (0.0) | 11 (0.1) | 0 (0.0) |  |
| Number of HLA-DR mismatches, n (%) | 1 (ref) | 64,865 (44.4) | 10,026 (42.0) | 54,839 (44.8) | <0.001 |
|  | 0 | 31,842 (21.8) | 6,022 (25.2) | 25,820 (21.1) |  |
|  | 2 | 49,537 (33.9) | 7,803 (32.7) | 41,734 (34.1) |  |
| not reported, n (%) |  | 27 (0.0) | 27 (0.1) | 0 (0.0) |  |
| Cold Ischemic time, mean (SD) | Hours | 18 (9.0) | 18.6 (8.7) | 17.9 (9.1) | <0.001 |
| not reported, n (%) |  | 6,906 (4.7) | 6,906 (28.9) | 0 (0.0) |  |
| ***Recipient factors*** |  |  |  |  |  |
| Age at Transplantation,  mean (SD) | Years | 53 (12.9) | 51.2 (12.4) | 53.4 (12.9) | <0.001 |
| Height, mean (SD) | Cm | 170.2 (10.8) | 170.3 (11.0) | 170.2 (10.7) | 0.023 |
| not reported, n (%) |  | 1,932 (1.3) | 1,932 (8.1) | 0 (0.0) |  |
| Weight, mean (SD) | Kg | 81.7 (19.4) | 78.7 (19.3) | 82.2 (19.3) | <0.001 |
| not reported, n (%) |  | 1,273 (0.9) | 1,273 (5.3) | 0 (0.0) |  |
| Dialysis duration, mean (SD) | Years | 3.9 (3.2) | 4.3 (3.6) | 3.8 (3.1) | <0.001 |
| cPRA, mean (SD) |  | 0.1 (0.3) | 0.1 (0.2) | 0.1 (0.3) | <0.001 |
| not reported, n (%) |  | 16,250 (11.1) | 16,250 (68.1) | 0 (0.0) |  |
| Race, n (%) | White (ref) | 84,689 (57.9) | 13,443 (56.3) | 71,246 (58.2) | <0.001 |
|  | Asian | 9,483 (6.5) | 1,477 (6.2) | 8,006 (6.5) |  |
|  | Black | 49,187 (33.6) | 8,513 (35.7) | 40,674 (33.2) |  |
|  | Multi | 485 (0.3) | 37 (0.2) | 448 (0.4) |  |
|  | Native | 1,685 (1.2) | 255 (1.1) | 1,430 (1.2) |  |
|  | Pacific | 742 (0.5) | 153 (0.6) | 589 (0.5) |  |
| Gender, n (%) | Female | 58,042 (39.7) | 9,511 (39.8) | 48,531 (39.7) | 0.608 |
| Diabetic, n (%) | Yes | 52,544 (35.9) | 6,776 (28.4) | 45,768 (37.4) | <0.001 |
| not reported, n (%) |  | 1,076 (0.7) | 1,076 (4.5) | 0 (0.0) |  |
| Hypertensive, n (%) | No (ref) | 13,539 (9.3) | 3,315 (13.9) | 10,224 (8.4) | <0.001 |
|  | Unknown | 19,550 (13.4) | 1,895 (7.9) | 17,655 (14.4) |  |
|  | Yes | 113,182 (77.4) | 18,668 (78.2) | 94,514 (77.2) |  |
| Pretransplant blood transfusions, n (%) | No (ref) | 79,606 (54.4) | 12,903 (54.0) | 66,703 (54.5) | <0.001 |
|  | Unknown | 43,276 (29.6) | 5,399 (22.6) | 37,877 (31.0) |  |
|  | Yes | 23,389 (16) | 5,576 (23.4) | 17,813 (14.6) |  |
| Angina, n (%) | No (ref) | 37,048 (25.3) | 15,449 (64.7) | 21,599 (17.7) | <0.001 |
|  | Unknown | 104,205 (71.2) | 6,537 (27.4) | 97,668 (79.8) |  |
|  | Yes | 5,018 (3.4) | 1,892 (7.9) | 3,126 (2.6) |  |
| Peripheral Vascular Disease,  n (%) | No (ref) | 132,028 (90.3) | 21,169 (88.7) | 110,859 (90.6) | <0.001 |
|  | Unknown | 4,779 (3.3) | 1,781 (7.5) | 2,998 (2.5) |  |
|  | Yes | 9,464 (6.5) | 928 (3.9) | 8,536 (7.0) |  |
| Drug Treated COPD, n (%) | No (ref) | 128,431 (87.8) | 21,590 (90.4) | 106,841 (87.3) | <0.001 |
|  | Unknown | 16,160 (11.1) | 2,079 (8.7) | 14,081 (11.5) |  |
|  | Yes | 1,680 (1.2) | 209 (0.9) | 1,471 (1.2) |  |
| Hepatitis C Virus Status, n (%) | No (ref) | 130,148 (89.0) | 19,544 (81.9) | 110,604 (90.4) | <0.001 |
|  | Unknown | 8,302 (5.7) | 3,015 (12.6) | 5,287 (4.3) |  |
|  | Positive | 7,821 (5.4) | 1,319 (5.5) | 6,502 (5.3) |  |
| ***Transplant outcomes*** |  |  |  |  |  |
| Allograft failure, n (%) | Yes | 25,655 (17.5) | 6,758 (28.3) | 18,897 (15.4) | <0.001 |
| Allograft survival time,  mean (SD) | Years | 4.1 (3.7) | 5.1 (4.3) | 3.7 (3.3) | <0.001 |
| Recipient death, n (%) | Yes | 28,940 (19.8) | 8,138 (34.1) | 20,802 (17.0) | <0.001 |
| Patient survival time,  mean (SD) | Years | 5.2 (3.9) | 6.5 (4.4) | 4.8 (3.6) | <0.001 |

SD = standard deviation; CVA = cerebrovascular accident; DCD = Donation after Circulatory Death; HLA = human leukocyte antigen; COPD = chronic obstructive pulmonary disease; cPRA = calculated Panel Reactive Antibodies

* ABO-compatible only

** p-value were computed with chi-squared test for discrete covariates and Kruskal-Wallis rank sum test for continuous covariates
